# Sex differences in elite track and field performances and inferences about steroid doping

**DOI:** 10.1101/2024.05.23.24307812

**Authors:** Diana B. Collantes, Jonathon W. Senefeld, Kathryn F. Larson, Doriane Lambelet Coleman, Michael J. Joyner, Shalaya Kipp

**Author notes:** **Correspondence** Shalaya Kipp, PhD, Department of Anesthesiology and Perioperative Medicine Mayo Clinic | 200 First Street SW | Rochester, MN 55905.

## Abstract

Females likely experience larger performance benefits from androgenic-anabolic steroids than males. We set out to determine if there were temporal differences in select athletics (track and field) records between females and males. Exploratory aims included: 1) evaluating the improvements in female and male world records over time, and 2) investigating the influence of doping programs on male and female world records before and after 1990 when sport’s governing bodies began to implement random out-of-competition and systematic in-competition drug testing. We collected the top 500 performances of all time for both sexes from an online database (worldathletics.org) in four running events (100m, 200m, 400m, and 800m) and two throwing events (discus throw and shot put). Data were stratified into quintiles based on world record ranking (1^st^ to 100^th^, 101^st^ to 200^th^, etc.). The temporal distribution of top 100 female performers was significantly earlier than top 100 male performers (year: 2000±1 vs 2005±1, respectively; p<0.001). Within event, the top performances occurred significantly earlier for females in the 800m (year: 1995±15 vs 2003±12; p<0.001) and shot put (year: 1992±14 vs 2003±17; p<0.001). Among females, world records rapidly improved through the 1980s, however following the 1990, the world records ceased to improve. Geographically, there was a greater representation of countries with state-sponsored doping programs, specifically among female performances. We postulate these sex differences in the temporal distribution of top performances are likely associated with enhanced effectiveness of exogenous androgens (steroid doping) among female athletes with lower endogenous androgen hormones compared to males.

**Highlights:** *What is the central question of this study?:* Despite a longstanding notion that top performance in athletics occurred earlier for females compared to males, likely due to the larger performance benefits of androgenic-anabolic steroids, no study has compared this temporal relation. Thus, we examined the temporal distribution of select athletics events between females and males.

*What is the main finding and its importance?:* The top-100 female performances occurred earlier than the top-100 male performances. The sex-related temporal differences were particularly notable for the shot put and 800m. Furthermore, there was a greater representation of countries with known state-sponsored doping programs, specifically among females. Our analyses suggest anabolic-androgenic steroids played a greater role in the world’s best female athletics performances.

## Introduction

Luminaries of physiology have long recognized the scientific value of ‘real world data.’ For example, Nobel Prize laureate A.V. Hill, explained that “in the study of the physiology of muscular exercise there is a vast store of accurate information, hitherto almost unexploited, in the records of athletic sports and racing” (Hill, 1925). Careful examination of elite performances can provide key insights into the limits of human performance and provide a testbed to draw inferential conclusions about the physiological determinants of human performance. Specifically, observational study and ‘passive experimentation’ of the world’s most elite athletes enable the study of rare human phenotypes exposed to extreme and prolonged exercise training that are logistically difficult to study outside of small case studies or series (Lucia et al., 2006; Robinson et al., 1937; Jones, 2006; Joyner et al., 2023).

Three fundamental observations about human performance provide a theoretical framework to consider world record performances. First, record performance velocities are slower with longer distances. The hyperbolic relation between average performance speed and the duration of the athletic event, first identified by A.V. Hill a century ago, is now generally referred to as the “power-duration relation” and is observed within a wide range of whole-body activities as well as exercise with a single muscle group (Hill, 1925; Poole et al., 2016; Burnley, 2009; Jones, 2010). Second, males outperform females by about 5 to 35% with variation depending on the physiological requirements of the athletic event (Hunter et al., 2023; Hunter, 2024). Third, record performances improve across time (Weiss et al., 2016). These improvements are attributed to the dissemination of empirical learning based on trial and error, more accessible science-based knowledge on best practices in training and competition (Sandbakk et al., 2023), advancements in sports technology or techniques (Dyer, 2015), and expanded competitive opportunities, particularly among females (Joyner et al., 2010).

World records that do not follow all three fundamental observations are suspected of representing performances enhanced by an unfair advantage to the athlete, such as performance-enhancing drugs (e.g., androgenic steroids or erythropoietin). The female athletics (track and field) world records that have failed to exhibit a consistent pattern of improvement over time have therefore aroused suspicions of doping. One example is the women’s world record in the 400 meters (47.60 s) set by Marita Koch of East Germany in 1985. Koch is widely believed to have used the anabolic steroid Turinabol for many years in the 1970s and 80s as part of a state-sponsored doping program (Franke & Berendonk, 1997).

Before sports governing bodies around the world committed to both random out-of-competition and systematic in-competition drug testing, robust state-sponsored and private-doping programs supported the widespread use by elite athletes of androgenic anabolic steroids among other performance enhancing drugs. Implementation of these doping programs occurred over a period of time, but the anti-doping commitment coalesced around 1990, in the period following the 1988 Olympic Games in Seoul. Therefore, we have broadly categorized two periods as pre and post 1990.

Androgenic-anabolic steroids are considered to be most effective among sporting events associated with maximal muscle power (Iyer and Handelsman, 2016). Additionally, they are likely more effective among females who have much lower endogenous androgen concentrations than males (Handelsman et al., 2018). Given the larger benefit females would experience from androgenic-anabolic steroids, and the historical, large-scale use of androgenic-anabolic steroids that resulted from state-sponsored and privately directed doping programs, we tested the hypothesis that top female performances would occur earlier than top male performances.

In a retrospective observational study, we evaluated performance year among the top 500 female and male athletes in events in the sport of athletics that may be influenced by the ergogenic effects of anabolic-androgenic steroids. Two additional exploratory aims of our analysis were to 1) evaluate the improvements in both female and male world records over time and 2) investigate the influence of state-sponsored doping programs by visually evaluating the geographical distribution of records in different periods of sport— before and after 1990.

## 2 Methods

### 2.1 Ethical approval

This retrospective observational study represents secondary use of data. All procedures accessed public information and did not require ethical review as determined by the Mayo Clinic Institutional Review Board in accordance with the Code of Federal Regulations, 45 CFR 46.102, and the Declaration of Helsinki. Performances of select outdoor athletics events of the top 500 male and female athletes were collected.

### 2.2 Primary analysis

World Athletics, the international governing body for the sport of athletics, maintains a database of the all-time world’s best performances for all its events. This publicly available online database (https://worldathletics.org/records/all-time-toplists/) served as the primary data source for this study. Data were extracted on 16 November 2023.

Performances from four track running events (100 m, 200 m, 400 m, and 800 m) and two field throwing events (discus throw and shot put) were collected, comprising the top 500 all-time best female and male performers (i.e., athletes may only be represented once in the database). Informed by *a priori* discussions, data associated with other athletics events were excluded from data collection and analysis for several reasons. First, given the preeminence associated with Olympic events, events not currently contested in the Olympics were excluded. Second, we excluded Olympic events that were not contested by both sexes in the pre and post 1990 testing periods (hammer throw, triple jump, 400m hurdles). Third, endurance events were excluded because they benefit differently from anabolic-androgenic steroid use (1500 m, 5000 m, 3000 m steeplechase, 10,000 m, marathon, and 20 km race walk). Fourth, several events were excluded because they have been associated with significant changes across time, specifically technique (high jump and long jump), equipment (javelin throw, pole vault), and competition length (110/100 m hurdles).

In this framework, up to 6,000 data points could be abstracted: 500 performances × 6 athletics events × 2 sexes. Although complete datasets were almost ubiquitously available, data associated with the discus throw and shot put were incomplete for female athletes. We included 467 performances for female athletes and 468 performers for male shot put athletes. Multiple male performers achieving the same mark (tie) accounted for the discrepancy in the number of performances. We included 351 performances for both female and male discus throwers. Thus, of the total 6,000 potential data points, 5,637 performances were included in analyses. Data extracted included: athlete’s name, nation represented, mark (finishing time or throw distance), and date of mark.

The primary outcome of these analyses was the year of competition associated with the top marks. Additionally, the data were stratified into quintiles based on performance place (i.e., 1^st^-100^th^; 101^st^-200^th^; 201^st^-300^th^; 301^st^-400^th^; 401^st^-500^th^) within each sex category (female, male).

### 2.3 Exploratory Analysis: World record progression

In a separate set of analyses to determine the progression of athletics world records across time, we generated a second database of all-time female and male records (i.e., individual athletes may be represented multiple times). This database comprised the top 2,000 all-time records from 1900 to 16 November 2023. To ensure earlier records were accounted for we included the top 2,000 all-time records from 1900 until 1980. The progression of world record was plotted by finding the top performance for every year.

### 2.4 Exploratory Analysis: Geographical representation of top athletes

To visually understand which countries contributed to the top performances, we created choropleth maps to depict the frequency of each over three different periods: (1) pre 1990 (2) post 1990 and (3) all-time performances (1950-2023).

Due to changes in nation state boundaries, some previous boundaries were recoded to their modern-day equivalent. Specifically: East Germany and West Germany were coded to Germany. The Soviet Union was coded to Armenia, Azerbaijan, Belarus, Estonia, Georgia, Kazakhstan, Kyrgyzstan, Latvia, Lithuania, Moldova, Russia, Tajikistan, Turkmenistan, Ukraine, and Uzbekistan. The Unified Team (1992 Olympic team for former Soviet athletes except those from the Baltics) was coded to Armenia, Belarus, Georgia, Kazakhstan, Russia, and Uzbekistan. Czechoslovakia was coded to Czech Republic and Slovakia. Yugoslavia was coded to Slovenia, Serbia, Montenegro, and Croatia. Former countries not included here did not have top athletes in the performance database.

### 2.5 Statistical analyses

We tested normality using a Shapiro-Wilk test. Because the distribution of performances across years was not normally distributed, we used Kruskal–Wallis ANOVAs to compare the differences between females and males within a given quintile. When significant, we used a Wilcoxon rank sum test to determine where the differences were (event). We applied Bonferroni correction for post hoc analysis to account for multiple comparisons and conservatively set our significance level to p<0.001. To test if the percentage of performances held before and after 1990 changed for a given country, we performed chi-squared tests. Analyses were performed using RStudio (Team, 2013; version 4.3.2, RStudio, Boston, MA). Data are reported as mean ± standard deviation in the tables and text, unless otherwise noted.

## 3 Results

### 3.1 Performance Ranking

Female top performances occurred significantly earlier than male top performances in the first, third and fourth quintiles (Figure 1, Table 1). When looking within each event, the top 100 female athletes (first quintile) for the 800m occurred significantly earlier than the top 100 male athletes for the 800m (year: 1995±15 vs 2003±12; P<0.001). The top female shot put performances also occurred earlier than the top male shot put performances in the first quintile (year: 1992±14 vs 2003±17; p<0.001), third quintile (year: 1998±16 vs 2006±13; p<0.001), and fourth quintile (year: 2000±17 vs 2007±15; p<0.001). Although female performances occurred earlier than the male performances in the other four events, we found no statically significant sex differences in the 100m, 200m, 400m or discus to conclude that females are likely more advantaged than males by steroid doping. Performances stratified by quintile, sex and event can be found in Table 1.

**Figure 1.**
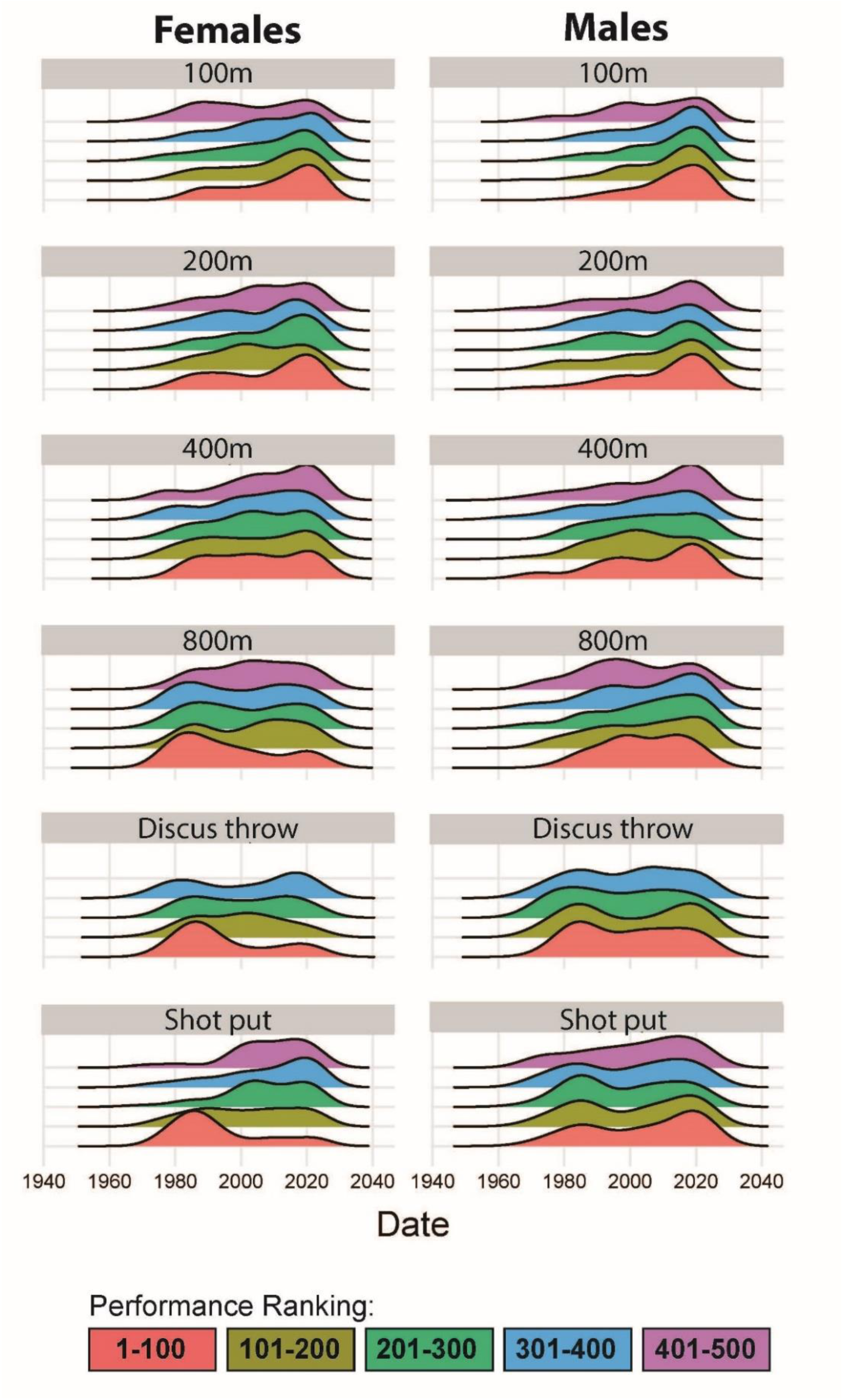
Temporal density plots of top performances for females and males athletics athletes. The 500 performances per event and sex are stratified by quintile according to ranking. The first quintile includes the top performance (1) to the 100^th^ performance. The second quintile includes the 101st performance to the 200^th^ performance. The third quintile includes the 201st performance to the 300^th^ performance. The fourth quintile includes the 301st performance to the 400^th^ performance. The fifth quintile includes the 401st performance to the 500th performance.

**Table 1.**
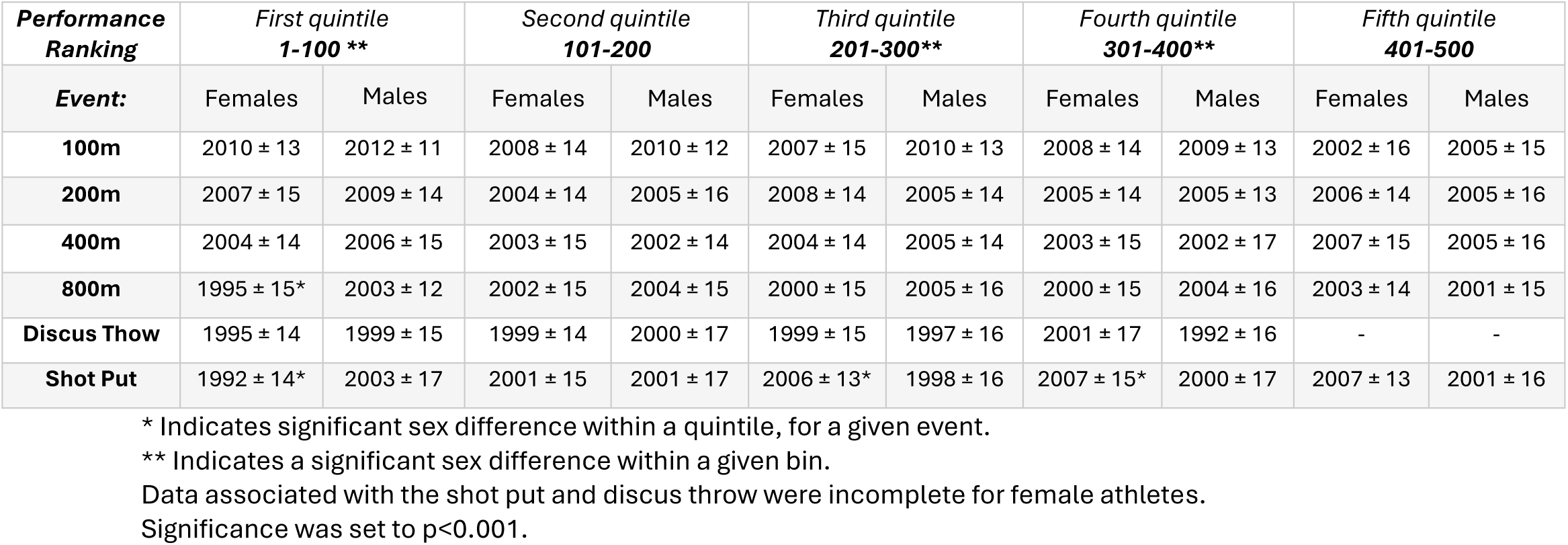
Mean year of top performances for a given ranking quintile. Data is presented mean ± SD.

### 3.2 Exploratory analysis of world record progression

We performed an exploratory analysis of the world record progression for select athletics events. There is a visible improvement in world record performances across all events and among both sexes until 1990 (Figure 2). Although world records associated with male athletes have continued to progress after 1990, world records among female athletes have not progressed since the 1980s. The mean world record year for female was 1987 while the mean world record year for males was 2009. Table 2 shows the year the current world records were set; notably, all female records in the studied events were set between 1983 and 1988.

**Figure 2.**
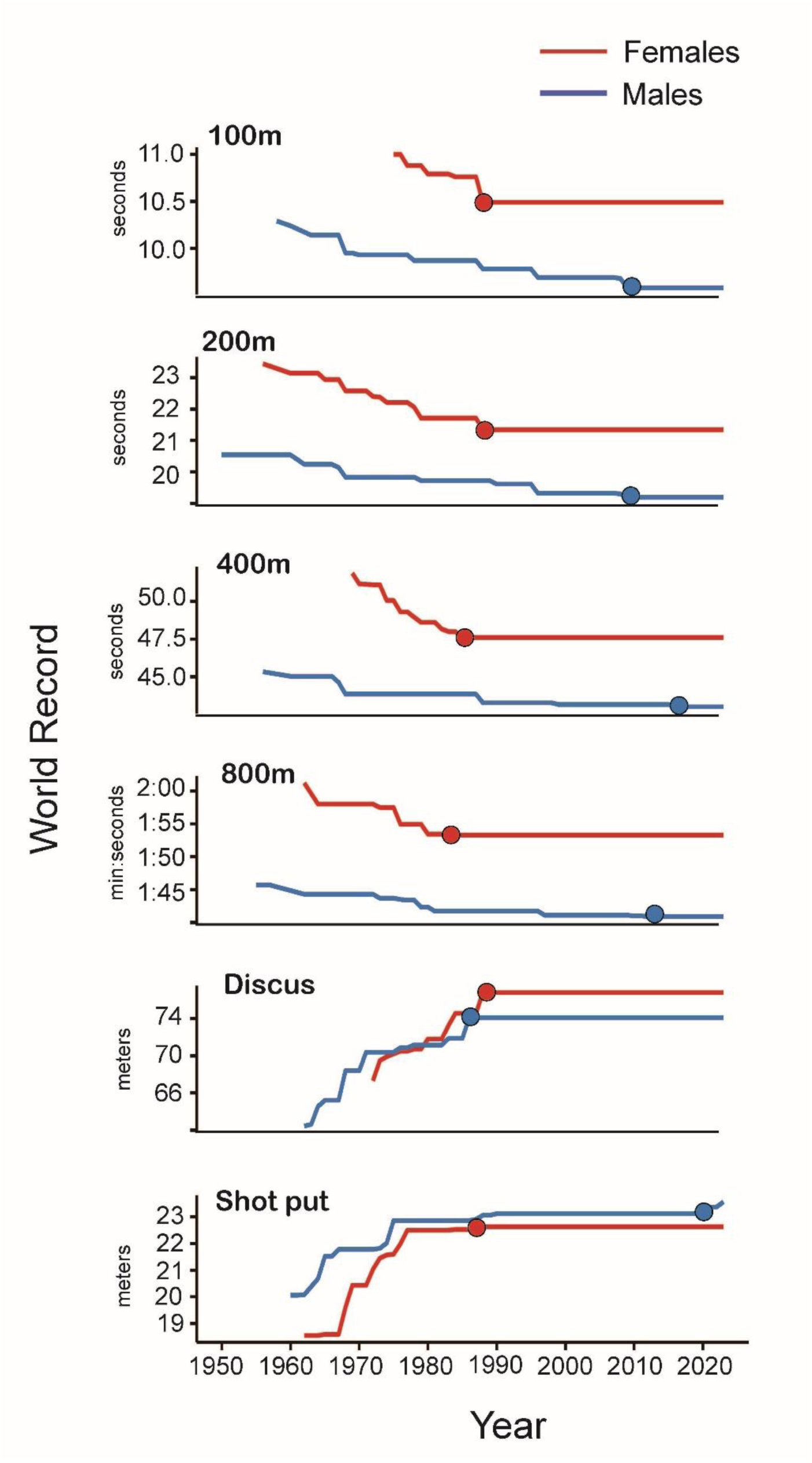
World record progression for Athletics events. Red lines represent female world records over time and blue lines represent male world records over time. The corresponding filled circles indicate current world records.

**Table 2.** Year associated with current world record ^a^.

| **Event** | **Females<sup>b</sup>** | **Males** |
| --- | --- | --- |
| 100 m dash | 1988 | 2009 |
| 200 m dash | 1988 | 2009 |
| 400 m dash | 1985 | 2016 |
| 800 m run | 1983 | 2012 |
| Discus throw | 1988 | 1986 |
| Shot put | 1987 | 2023 |

<sup>a</sup> Current world records as of 25 March 2024.
<sup>b</sup> Note each of the selected female world records were set in the 1980s before ‘out-of-competition’ drug testing was implemented in 1989.

### 3.3 Exploratory geographical analysis

We performed an exploratory analysis of the geographical representation of top performances (Figure 3 and Table 3), in part to investigate the prevalence of top performances from countries with documented state-sponsored doping programs, specifically the German Democratic Republic (Fitch 2008; Franke & Berendonk) and former Soviet Union (Kalinski & Kerner, 2002). Among these formerly recognized regional areas (German Democratic Republic and Soviet Union) and their modern-day equivalents (Germany and namely Russia, respectively), there were markedly more performances before 1990. While the Soviet Union was comprised of many countries, we have chosen to focus on Russia given the large number of performances they achieved in the post 1990 period. We calculated that 23% and 10.99% of the top all-time performances up to 1990 were female and male athletes from Germany, respectively. After 1990, there was a significant decrease in the records held by German female (4.28%, p<0.001; *X^2^* =164.32) and male athletes (2.8%, p<0.001; *X^2^* =65.33). Russia showed a similar pattern, accounting for 34.08% and 14.13% of the top female and male all-time performance up to 1990. After 1990, there was a significant decrease in the performances held by Russian females (8.74%, p<0.001; *X^2^* =179.02) and male athletes (1.42%, p<0.001; *X^2^* =167.14). Other countries that held over 3% of the total performances before 1990 included: Romania, Bulgaria, the United Kingdom, and the United States. The United States recorded a large fraction of the male records prior to 1990 (37.36%), however in contrast to Germany and Russia, the fraction of top records by male athletes in the United States was maintained after 1990 (31.26%, p=0.04; *X^2^* =4.15). Females from United States represented 10.38% of the top female athletes up to 1990 and increased to 29.69% from 1990 to 2023 (p<0.001; *X^2^*=69.50), a change which may be attributed to more competitive opportunities, professionalization for females and increased worldwide doping control.

**Figure 3.**
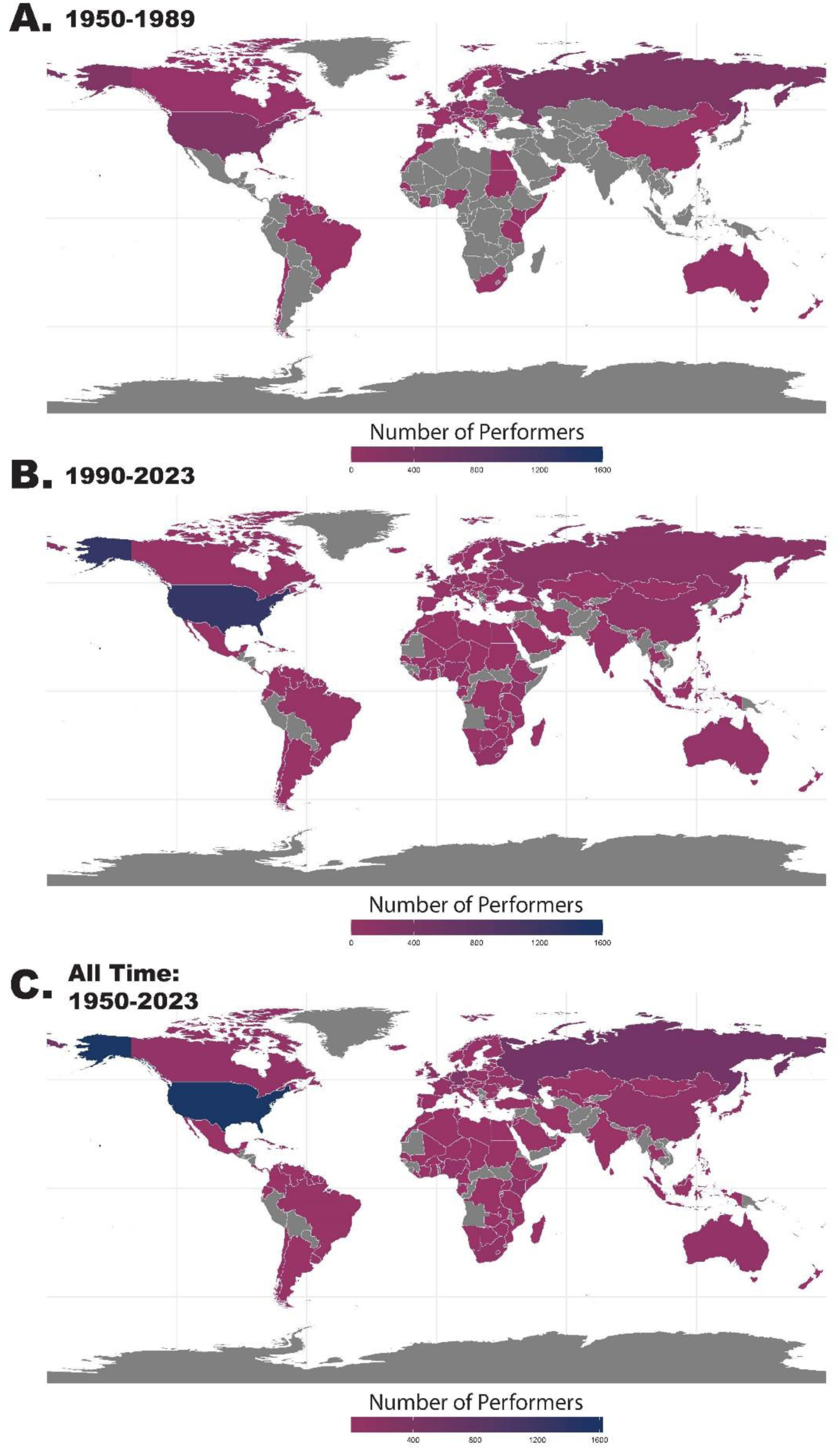
World Map showing distribution of top athletics athletes. The top panel (A) shows the number of athletes from each country from 1950-1989 (year out of competition drug testing started). The middle panel (B) shows the number of athletes from 1990-2023. The bottom panel (C) shows the all-time number of athletes (1950-2023).

**Table 3.** Percentage of performances held by female and male athletes for their respective counties from 1950-1989, before the implementation of out-of-competition drug testing; and from 1990-2023, after the implementation of out-of-competition drug testing. We included only countries that held >3% of the records for either sex at any time point.

| **Country:** | **1950-1989**<br>Before implementation of out-of-competition testing |  | **1990-2023**<br>After implementation of out-of-competition testing |  | *Female* |  | *Male* |  |
| --- | --- | --- | --- | --- | --- | --- | --- | --- |
| | Females | Males | Females | Males | $\chi^2$ | p-value | $\chi^2$ | p-value |
| **United States** | 10.4% | 37.4% | 29.7% | 31.3% | 69.5 | <0.001* | 4.2 | 0.042 |
| **Russia** | 34.1% | 14.1% | 8.7% | 1.4% | 179.0 | <0.001* | 167.1 | <0.001* |
| **Germany** | 23.0% | 11% | 4.3% | 2.8% | 164.3 | <0.001* | 65.3 | <0.001* |
| **China** | 1.0% | 0% | 5.8% | 0.6% | 26.0 | <0.001* | - | - |
| **United Kingdom** | 3.8% | 3.1% | 3.7% | 4.8% | <0.01 | 0.910 | 2.9 | 0.090 |
| **Romania** | 4.9% | 0.3% | 0.9% | 0.3% | 45.0 | 0.001* | 0.01 | 0.978 |
| **Jamaica** | 0.8% | 1.6% | 6.5% | 5.7% | 32.9 | 0.001* | 17.4 | 0.003 |
| **Kenya** | 0.1% | 2.8% | 1.1% | 5.3% | 5.3 | 0.210 | 6.2 | 0.012 |
| **Bulgaria** | 4.6% | 1.9% | 0.7% | 0.2% | 48.6 | 0.001* | 24.8 | <0.001* |

\*Significance was set to $p < 0.001$ .

## 4 Discussion

This retrospective, observational study used ‘real-world data’ – elite athletics performances of females and males – to explore the temporal and geographical distributions of top athletes in select events that may have been influenced by the ergogenic effects of anabolic-androgenic steroids. Available data suggest an influential role of anabolic-androgenic steroids on the world’s best athletics athletes, particularly among female athletes. Several different analyses supported this primary finding. First, among the top 100 best athletics athletes within select events, the performance year associated with top athletes occurred earlier for females compared to males. These sex-related temporal differences were particularly notable for two events—shot put and 800s. Second, world records did not progress after 1990 among females. Lastly, there was a greater representation of countries with known state-sponsored doping programs, specifically among female performances. In this context, our analyses suggest anabolic-androgenic steroids played a substantial role in the world’s best athletics performances, especially among female athletes.

### 4.1 Potential contribution of anabolic-androgenic steroids

Androgenic-anabolic steroids, synthetic derivatives of the hormone testosterone, have long been recognized for their potential to enhance athletic performance and have influenced sporting culture particularly in countries with state-sponsored doping programs (Handelsman, 2021). Notably, early scientists and physicians found that androgen doping was particularly effective in females, especially in events requiring speed and strength (Franke & Berendonk,1997). Because adult males have about 15-fold greater endogenous testosterone concentrations than adult females (Senefeld & Hunter, 2024; Nokoff et al., 2023), the sigmoidal dose response relationship for exogenous androgens differs between females and males (Huang & Basaria, 2018; Handelsman et al., 2018). Females with lower endogenous testosterone concentrations likely experience greater performance benefits with exposure to lower doses of exogenous androgens compared to males who have higher endogenous testosterone concentrations (Huang & Basaria, 2018; Handelsman et al., 2018). In the dose and response curve context, with a similar exposure to exogenous androgens, females may experience substantial performance enhancement and males likely experience a small, blunted ergogenic effect compared to females.

### 4.2 Historical context

The androgenic effects associated with testosterone were characterized in the 1930s and exogenous androgens were adopted in sport during the early years of the Cold War likely due to sociopolitical pressures to demonstrate geopolitical supremacy (Handelsman, 2021; Iyer and Handelsman, 2016). In this framework, several countries embraced a national system of androgen doping to improve sport performance commonly known as ‘state-sponsored doping programs’ (Fitch, 2008). The differential performance enhancing effects of androgen doping on females was noted. For example, after the collapse of the German Democratic Republic, documents were recovered that provided detailed information of the specifics of their doping program, notably the heavy use of androgenic-anabolic steroids on female athletes:

> *“From our experiences made so far it can be concluded that women have the greatest advantage from treatments with anabolic hormones with respect to their performance in sport*….”-- March 3, 1977,SMD Deputy Director Hoppner; Vol. II of his Stasi reports under the code name “Technik,” pp. 243–44. (Franke & Berendonk, 1997).

Although the International Olympic Committee prohibited the use of androgenic anabolic steroids in-competition in 1974, thereafter drug testing at the Olympics was conducted in an overt and predictable pattern allowing the opportunity for athletes temporarily to cease androgen use before the Games. In this context, widespread androgenic anabolic steroid use continued until the sport’s governing bodies became serious about comprehensive in and out of competition drug testing in the period following the 1988 Olympic Games in Seoul (Blackwell, 1991; Jacobs & Samuels, 1994). This shift in policy together with the geopolitical events that caused the collapse of the German Democratic Republic, the Soviet Union, and their respective doping programs is associated with a stagnation in female records.

It is noteworthy that in the West, societal norms around female participation in sports changed, opportunities for participation expanded, and participation numbers surged in the post 1990 period. In light of these developments, Whipp and Ward (1992) calculated a steeper progression of the female world records for track events, extrapolating mean running velocities to pose the hypothetical question ‘*will women soon outrun men*?” and it was postulated that female world records would have declined to a greater extent than male records in the past three decades (Cheuvront et al., 2005). However, this was not observed and, as illustrated in the current study, it contrasts with the patterns observed in world record progression for select athletics events.

### 4.3 Perspective

Careful examination of elite athletics data can provide key insights into human performance and provide inferential conclusions about performance physiology. Although our study faced several limitations enumerated below, our results provide several lines of evidence suggesting an influential role of anabolic-androgenic steroids on world’s best athletics performances, particularly among female athletes competing in 800s and shot put. First, female top performances occurred earlier than male top performances. Second, our exploratory analyses of world record progression showed female records were established in the 1980 and 1990, while male records continue to improve. Third, geographic analysis reveals countries with state-sponsored doping programs are over-represented prior to 1990.

### 4.4 Limitations

Our study quantified records of select athletics events without assessment of physiologic factors and other influential factors, such as prize money incentives, better tracks, and advanced footwear technology (Dyer, 2015). We were limited to publicly available data that did not include information on performance-influencing factors such as performance enhancing drug use. While anabolic-androgenic steroids have been the focus of this paper, other performance enhancing drugs may have been taken concurrently, such as human growth hormone. Another potential limitation was that our geographic analysis used the country an athlete represented, which may not have reflected where the athlete was from or trained. Also, our analyses were restricted to a subset of events, which limits the generalizability of our findings. In this framework, our retrospective, observational study design limits the physiologic and mechanistic insights that can be deduced directly from the available data. Despite these limitations, our study provides several different analyses which converge in the inferential conclusion that anabolic-androgenic steroids have influenced the world’s best performances and world records in athletics, particularly among female athletes.

## 5 Conclusions

Our analyses show that among the top 100 best athletics athletes in several select events, the performance year associated with top performances was earlier for females compared to males. Furthermore, the female world records in each of these events were set in the 1980s, while male world records continue to improve. Exploratory geographic analysis also revealed that countries which had state-sponsored doping programs at the time are over-represented prior to implementation of systematic in- and out-of-competition drug testing, the pre 1990 period. These analyses suggest an influential role of anabolic-androgenic steroids on world’s best athletics performances, particularly among female athletes competing in 800m and shot put events.

## Author Contributions

Conceptualization: Jonathon W. Senefeld, Michael J. Joyner, Shalaya Kipp. Data curation: Diana B. Collantes, Jonathon W. Senefeld. Kathryn F. Larson, and Shalaya Kipp. Data analysis: Shalaya Kipp. Writing—original draft: Diana B. Collantes and Shalaya Kipp. Writing—review & editing: Diana B. Collantes, Jonathon W. Senefeld, Kathryn F. Larson, Doriane Lambelet Coleman, Michael J. Joyner, and Shalaya Kipp. All authors have read and approved the final version of the manuscript and agree to be accountable for all aspects of the work in ensuring that questions related to the accuracy or integrity of any part of the work are appropriately investigated and resolved. All persons designated as authors qualify for authorship, and all those who qualify for authorship are listed.

## Data Availability Statement

The analyzed datasets supporting this study are publicly available online at the World Athletics website. Supporting data are available upon reasonable request.

